# Distributional Diagnosis and Calibration with Negative Controls for Outcome-wide Real-world Evidence

**DOI:** 10.64898/2026.07.08.26357550

**Authors:** Huiyuan Wang, Bingyu Zhang, Yuqing Lei, Yiwen Lu, Dazheng Zhang, Xinyao Jian, Yuru Zhu, Wenjie Hu, Haitao Chu, Yong Chen, Marc A. Suchard, Patrick B. Ryan, George Hripcsak, David A. Asch, Yun Lu, Bin Yu, Martijn J. Schuemie, Yumou Qiu, Yong Chen

## Abstract

Glucagon-like peptide-1 receptor agonists (GLP-1RAs) have been linked to heterogeneous, potentially pleiotropic effects across organ systems, motivating outcome-wide comparative risk profiling in real-world data. A central challenge in such analyses is *residual bias* that remains after adjustment for observed confounders, which can distort effect estimates and mis-calibrate uncertainty. We present distributional diagnosis and calibration (DC), which uses panels of negative control outcomes (NCOs) to diagnose residual bias and calibrate uncertainty. DC evaluates null behavior via *p*-value uniformity and empirical coverage across NCOs, and uses the empirical distribution of NCO effect estimates to calibrate confidence intervals for prespecified primary outcomes. DC is modular: it can wrap around commonly used causal inference methods and operates directly on summary statistics, supporting collaborative research under data-sharing constraints. Using electronic health records from a large U.S. clinical research network (152.7 million patients), we compared GLP-1RAs with sodium–glucose cotransporter 2 inhibitors across 15 prespecified outcomes spanning cardiovascular, mental health, and genitourinary domains using four causal estimators. Across outcomes and methods, DC diagnostics revealed substantial and method-dependent residual systematic error. DC calibration attenuated systematic error signals observed in negative controls and yielded more stable, better-calibrated estimates for clinical outcomes, supporting DC as a practical strategy to strengthen the credibility of outcome-wide real-world CER.

**Disclaimer:** The contents are solely the responsibility of the authors and do not necessarily represent the official views of, or an endorsement by, Food and Drug Administration (FDA)/Department of Health and Human Services (HHS) or the U.S. Government.

## Introduction

GLP-1 receptor agonists (GLP-1RAs) have reshaped the clinical landscape of type 2 diabetes management. Although initially developed for glycemic control, GLP-1RAs such as semaglutide have demonstrated substantial benefits for weight loss and cardiovascular risk reduction [1]. However, real-world adoption has outpaced the evidence base from randomized controlled trials (RCTs), particularly for secondary outcomes and for patients who differ from trial populations in comorbidity burden, healthcare access, and prescribing context [2, 3].

Observational studies further suggest a pleiotropic profile of GLP-1RAs, with reported associations spanning multiple organ systems, from gastrointestinal adverse events to potential protective signals for substance use disorders and depression [4–6]. This breadth highlights the need to characterize outcome-wide risk–benefit profiles in routine care, where treatment decisions increasingly require balancing effects across cardiovascular, metabolic, neuropsychiatric, and other domains. Large-scale real-world data resources, including electronic health records (EHR) and claims networks, make such systematic evaluation feasible. Yet the same setting poses a distinct inferential challenge: when many outcomes are examined simultaneously, unmeasured confounders can generate correlated spurious signals across the phenome. For example, GLP-1RA users may appear healthier because of differential healthcare engagement, or sicker because of indication-driven prescribing; either pattern can shift multiple endpoints and blur causal effects with systematic bias. This setting motivates a general need for scalable, data-driven tools to diagnose and calibrate residual systematic error in outcome-wide real-world evidence generation.

A fundamental barrier is that counterfactual outcomes are unobserved, so the magnitude and structure of bias cannot be measured directly from the primary endpoint alone. Negative control outcomes (NCOs) — outcomes known not to be causally affected by the treatment — provide a practical lens for detecting hidden bias: any apparent treatment effect on an NCO reflects residual systematic error [7, 8]. Despite their appeal, negative controls are historically used as informal checks [9]. More recent methods that incorporate negative controls into causal estimation often rely on strong structural assumptions or causal restrictions that can be difficult to justify in complex real-world settings [10–12]. Moreover, most of these approaches focus on bias correction but offer limited tools to assess whether meaningful residual bias persists after adjustment, complicating interpretation of real-world evidence. There remains a need for principled, data-driven approaches that leverage *panels* of NCOs to characterize systematic error and support calibrated inference for outcomes whose true effects are unknown.

Here we introduce *distributional diagnosis and calibration* (DC), a unified framework that leverages a panel of NCOs to model and adjust for systematic error in non-interventional studies. Rather than requiring detailed specification of causal structure, DC attributes systematic components of estimation error across outcomes to shared features of the data environment, study design, and analytic pipeline. The key premise is a form of *bias exchangeability*: when the primary outcome and NCOs are analyzed within the same data source using standardized cohort construction, covariate definitions, and a common estimation procedure, residual confounding and model misspecification tend to induce systematic error components with similar behavior across outcomes. By learning the empirical distribution of estimated NCO effects, DC provides an internal diagnostic of residual bias and yields calibrated uncertainty for the primary endpoint.

We demonstrate DC using data from the TriNetX Research Network, a global EHR platform aggregating de-identified patient data from hundreds of healthcare organizations. To characterize residual bias in this setting, we leverage a prespecified panel of 95 NCOs expected to have null treatment effects based on clinical knowledge. We perform leave-one-out analyses, treating each NCO in turn as a pseudo-primary endpoint, to map patterns of spurious association and quantify the magnitude and dispersion of systematic error under known null conditions. Guided by these empirically observed bias patterns, we estimate and calibrate comparative treatment effects of GLP-1RAs versus sodium–glucose cotransporter 2 inhibitors (SGLT2is) across the 15 prespecified outcomes. Together, these results illustrate how DC provides an internal, data-driven assessment of residual systematic error and yields calibrated uncertainty for outcome-wide real-world comparative effectiveness analyses.

## Results

### Overview of DC

Figure 1 summarizes distributional diagnosis and calibration (DC), a post-estimation module for diagnosing and calibrating residual systematic error in observational causal analyses. DC takes as input the point estimates and standard errors produced by a prespecified estimation pipeline and returns (i) diagnostics that characterize whether negative controls behave as expected under the null and (ii) calibrated inference for the prespecified clinical outcomes. Conceptually, DC consists of a setup step, a diagnostic layer based on a negative-control panel, and a calibration step that propagates empirically observed systematic error into the primary inference.

**Figure 1.**
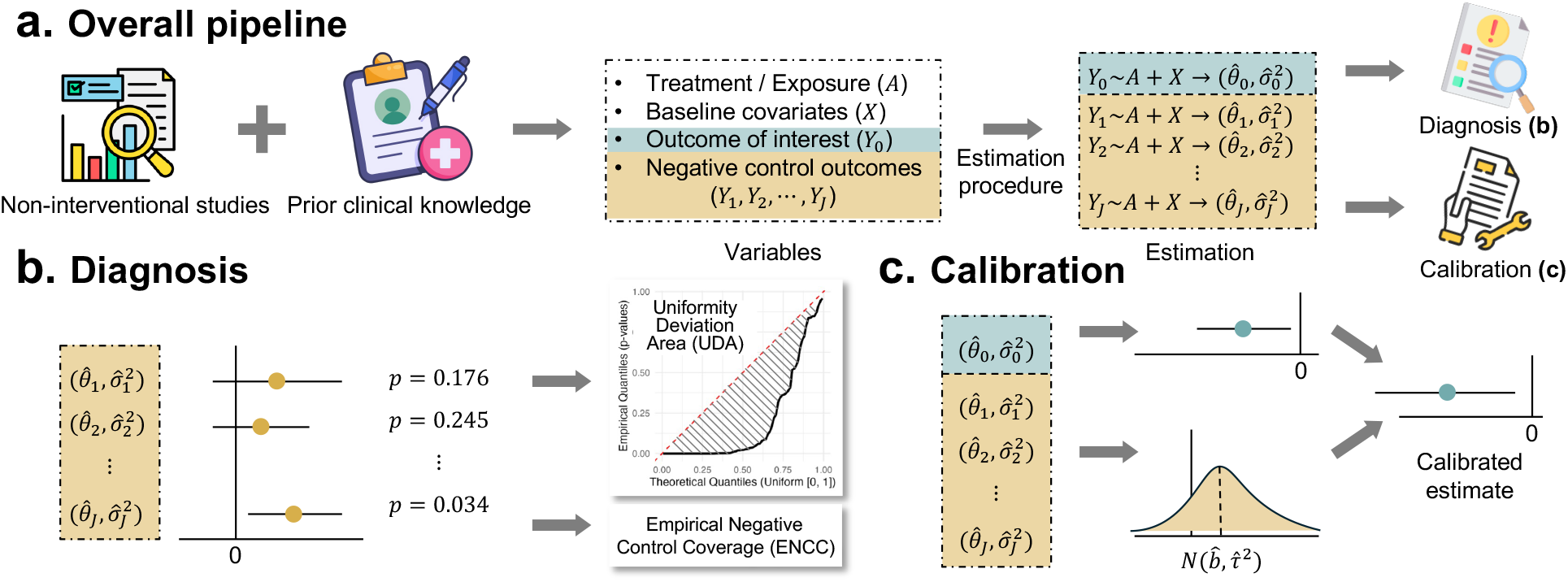
Overview of distributional diagnosis and calibration using negative control outcomes. **a.** Study setup: integrate real-world data and prior clinical knowledge to define treatment (*A*), covariates (*X*), primary outcome (*Y*_0_), and a prespecified panel of negative control outcomes (*Y*_1_, …, *Y*_*J*_), followed by model-based estimation. **b**. Diagnosis: use the negative-control panel to characterize residual systematic error (e.g., QQ plots and empirical null coverage). **c**. Calibration: use the empirical distribution of negative-control estimates to calibrate the primary estimate and its uncertainty.

#### Diagnostic layer (b)

DC uses the negative-control panel as an exploratory lens to map residual bias patterns induced by the shared data source and analytic pipeline. First, we examine quantile–quantile (QQ) plots of negative-control *p*-values. If residual systematic error is negligible and uncertainty is well-calibrated among the negative control outcomes whose true treatment effects are expected to be null, the *p*-values should be approximately uniform and the QQ curve should track the 45-degree reference line; systematic departures suggest residual confounding and/or mis-calibrated uncertainty. We summarize the deviation using the Uniformity Deviation Area (UDA), defined as the area between the empirical QQ curve and the 45-degree line, with larger values indicating stronger evidence of systematic error. Second, Empirical Negative Control Coverage (ENCC) quantifies how often nominal confidence intervals for negative controls include zero. Coverage below the nominal level indicates remaining bias and/or underestimated uncertainty.

#### Calibration layer (c)

DC calibrates inference using the empirical distribution of negative-control estimates. In brief, DC recenters the primary estimate by the average estimated negative-control effect and inflates uncertainty to reflect dispersion across negative controls, thereby accounting for both systematic shift and over/under-dispersion. This calibration is adaptive: when negative controls show little evidence of systematic error, DC makes minimal changes; when negative controls indicate substantial residual bias, DC applies a stronger correction and widens uncertainty accordingly. Under mild regularity conditions, the resulting confidence intervals achieve valid coverage.

#### Application to outcome-wide comparative effectiveness in TriNetX

We applied DC to the TriNetX Research Network, a global EHR platform aggregating de-identified patient data from hundreds of healthcare organizations. The clinical objective was to compare GLP-1RAs versus SGLT2is across 15 prespecified outcomes spanning cardiovascular, mental health, and genitourinary domains. Each outcome was analyzed as an incident event using an outcome-specific washout procedure, yielding outcome-specific analytic cohorts. We implemented DC with four causal estimators and applied a common prespecified negative-control panel to characterize residual systematic error in this data environment.

Across the 15 outcomes, DC diagnostics provided an internal assessment of residual systematic error under each estimation pipeline, and DC calibration yielded calibrated uncertainty for comparative effect estimates. In the main text, we report the calibrated outcome-wide results and the corresponding negative-control diagnostics; additional results under alternative estimators and extended analyses are provided in the *Supplementary Information*.

### DC diagnostics reveal residual systematic error across estimation methods

We assessed residual systematic error across combinations of estimation method and covariate adjustment. In each setting, we applied DC diagnostics to 95 prespecified NCOs, which are expected to have null treatment effects. This design provides an internal check of whether the analysis pipeline yields well-calibrated null behavior in the same data environment as the clinical outcomes.

#### QQ plots and UDA for uncalibrated estimates

Figure 2 shows substantial departures from Uniform(0, 1) behavior for uncalibrated NCO *p*-values (gray curves) across estimation methods and adjustment levels, indicating residual systematic error and/or mis-calibrated uncertainty that persists after standard confounding adjustment. Increasing covariate adjustment generally moved the *p*-value distributions closer to the 45-degree line, but noticeable deviations remained even under extensive adjustment (up to *d* = 178 covariates), suggesting that richer adjustment alone did not fully restore well-calibrated null behavior.

**Figure 2.**
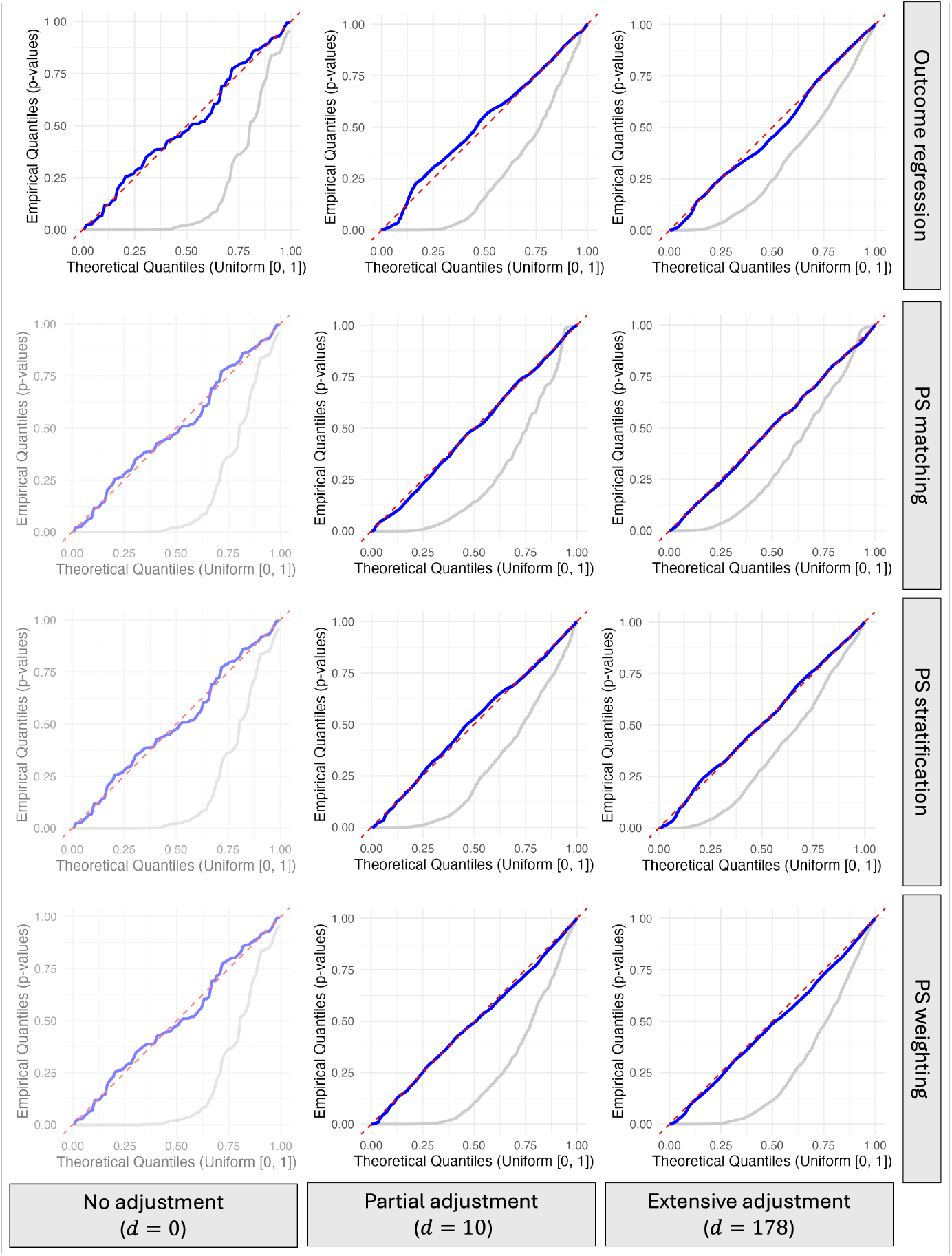
DC diagnostics before and after calibration. Each panel shows a QQ plot comparing empirical *p*-values from 95 NCOs to the Uniform(0, 1) reference across covariate adjustment levels (columns) and estimation methods (rows). Gray curves show uncalibrated *p*-values; blue curves show calibrated *p*-values obtained by leave-one-out DC using the NCO panel. The number of adjusted covariates is denoted by *d* (the no-adjustment column is identical across methods). The red dashed line indicates ideal uniformity.

To summarize these departures, we computed the Uniformity Deviation Area (UDA). Table 1 reports UDA values together with empirical null quantiles. For each estimator, UDA decreases systematically with more extensive covariate adjustment, indicating improved (but still imperfect) alignment with the expected null behavior. The Q95 and Q99 thresholds are obtained from a parametric bootstrap null that preserves the observed covariate structure by re-simulating treatment and outcomes from fitted models (excluding unmeasured confounding under the null); implementation details are provided in *Methods*.

**Table 1:** Quantitative diagnostics for *p*-value uniformity. We report UDA across estimation methods and covariate adjustment levels, before and after DC calibration. For each setting, Q95 and Q99 denote the 95th and 99th percentiles of the UDA obtained by a parametric bootstrap procedure; UDA values exceeding these thresholds indicate departures from Uniform(0, 1) behavior. Across settings, uncalibrated UDAs substantially exceeded the empirical null thresholds, whereas DC calibration consistently reduced UDA below the corresponding Q95 benchmark, indicating markedly improved alignment with the expected null behavior.

| Adjustment Level | Method | Calibration | Area | Q95 | Q99 |
| --- | --- | --- | --- | --- | --- |
| No adjustment ($d = 0$) | None (Unadjusted) | Uncalibrated | 0.303 | 0.062 | 0.073 |
|  |  | Calibrated | 0.023 | 0.062 | 0.073 |
| Partial adjustment ($d = 10$) | PS matching | Uncalibrated | 0.225 | 0.075 | 0.092 |
|  |  | Calibrated | 0.028 | 0.075 | 0.092 |
|  | PS stratification | Uncalibrated | 0.226 | 0.083 | 0.109 |
|  |  | Calibrated | 0.020 | 0.083 | 0.109 |
|  | PS weighting | Uncalibrated | 0.251 | 0.080 | 0.104 |
|  |  | Calibrated | 0.016 | 0.080 | 0.104 |
|  | Outcome regression | Uncalibrated | 0.245 | 0.137 | 0.154 |
|  |  | Calibrated | 0.027 | 0.137 | 0.154 |
| Extensive adjustment ($d = 178$) | PS matching | Uncalibrated | 0.181 | 0.075 | 0.095 |
|  |  | Calibrated | 0.022 | 0.075 | 0.095 |
|  | PS stratification | Uncalibrated | 0.161 | 0.072 | 0.091 |
|  |  | Calibrated | 0.014 | 0.072 | 0.091 |
|  | PS weighting | Uncalibrated | 0.232 | 0.082 | 0.104 |
|  |  | Calibrated | 0.021 | 0.082 | 0.104 |
|  | Outcome regression | Uncalibrated | 0.180 | 0.090 | 0.110 |
|  |  | Calibrated | 0.027 | 0.090 | 0.110 |

#### Empirical coverage for uncalibrated estimates

We further assessed uncertainty calibration by examining the empirical coverage of nominal 95% confidence intervals across the 95 NCOs. Specifically, we drew 200 random subsamples from the analytic cohort; within each subsample, we applied the prespecified causal estimator to each of the 95 NCOs to obtain a nominal 95% confidence interval for its estimated treatment effect, and then computed the fraction of these intervals that contained zero. This yields one empirical coverage value per subsample. Figure 3 summarizes the resulting distribution of coverage values across subsamples.

**Figure 3.**
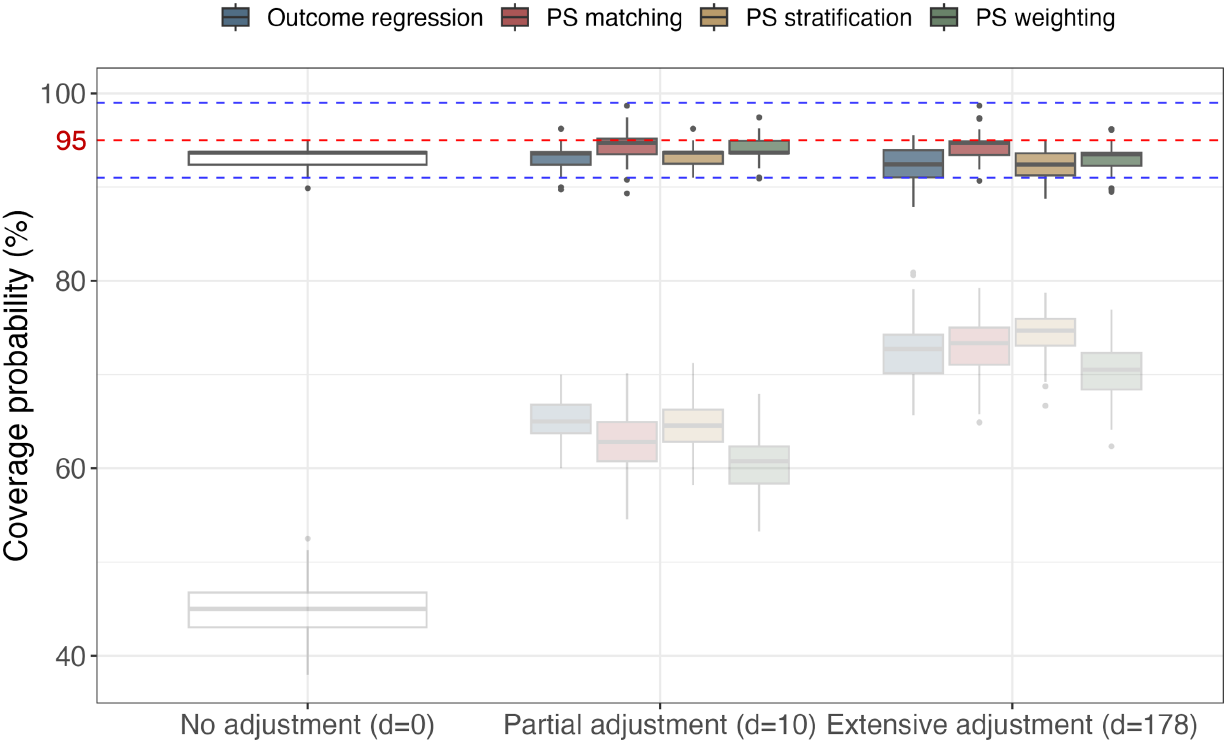
Empirical negative-control coverage before and after calibration. For each subsample and analysis setting, we applied LOO-based DC calibration to obtain calibrated 95% confidence intervals for the treatment effects of all NCOs, and computed the fraction of NCO intervals that contained zero (one coverage value per subsample). Boxplots summarize coverage across 200 random subsamples, stratified by covariate adjustment level (x-axis) and estimation method (color). The red dashed line marks nominal 95% coverage; blue dashed lines indicate 91% and 99% reference bounds. Faint boxplots show uncalibrated coverage for comparison (reproduced from Fig. S.3 in the Supplementary Material).

Across estimation methods and covariate adjustment levels, uncalibrated procedures (faint gray boxplots) frequently fell below the nominal 95% level, indicating mis-calibrated uncertainty and/or residual systematic error. Although more extensive covariate adjustment tended to improve coverage on average, coverage remained variable even under extensive adjustment, reflecting sensitivity to sampling variation and persistent systematic error in the observational pipeline.

Together with the QQ-based diagnostics, these results show that standard covariate adjustment alone often does not restore well-calibrated null behavior, motivating post-estimation calibration.

### DC calibration restores well-calibrated null behavior across estimation methods

We next evaluated whether DC yields well-calibrated confidence intervals and hypothesis tests in the presence of residual systematic error. Because the same negative-control panel informs both diagnosis and calibration, we used a leave-one-out (LOO) strategy to avoid reusing the same NCO for fitting and evaluation. In each iteration, one NCO was held out, the remaining NCOs were used to estimate the empirical bias distribution, and the held-out NCO was then calibrated using this distribution (see *Methods*).

#### Empirical coverage after calibration

To assess stability across sampling variation, we repeated the LOO procedure across 200 random subsamples and computed an empirical null-coverage value in each subsample. Figure 3 shows that DC calibration brings empirical coverage closer to the nominal 95% level across estimation methods and adjustment levels, while substantially reducing between-subsample variability relative to uncalibrated analyses. The largest improvements occur in under-adjusted settings, consistent with DC correcting both systematic shift and underestimation of uncertainty under residual bias.

#### Consistency with QQ-based diagnostics

The coverage improvements are consistent with the QQ-based diagnostics: in Fig. 2, LOO-based calibration moves negative-control *p*-value distributions closer to Uniform(0, 1) across methods and adjustment levels. Likewise, Table 1 shows that calibrated UDA values fall below the corresponding empirical null Q95 thresholds across settings, indicating markedly improved null behavior after calibration.

### DC calibration gains efficiency with stronger covariate adjustment

Having established that DC improves uncertainty calibration under residual systematic error, we next examined how covariate adjustment affects the precision of calibrated inference. DC widens confidence intervals to reflect uncertainty induced by residual systematic error estimated from the negative-control panel, which raises a practical question: can stronger covariate adjustment reduce the amount of inflation required for calibration?

To address this question, we applied LOO-based DC to the 95 NCOs under increasing levels of covariate adjustment while holding the estimator fixed. At each adjustment level, we computed the mean length of calibrated and uncalibrated 95% confidence intervals across NCOs and summarized results over 200 random subsamples. We report results for propensity score stratification in the main text; results for propensity score weighting, propensity score matching, and outcome regression are provided in the *Supplementary Materials*.

Figure 4 shows a clear efficiency gain with stronger adjustment. As more covariates are included, calibrated intervals become shorter on average, and the ratio of calibrated to uncalibrated interval lengths decreases, indicating that the additional uncertainty propagated by DC is substantially reduced under richer adjustment. At the same time, empirical null coverage remains close to the nominal 95% level after calibration.

**Figure 4.**
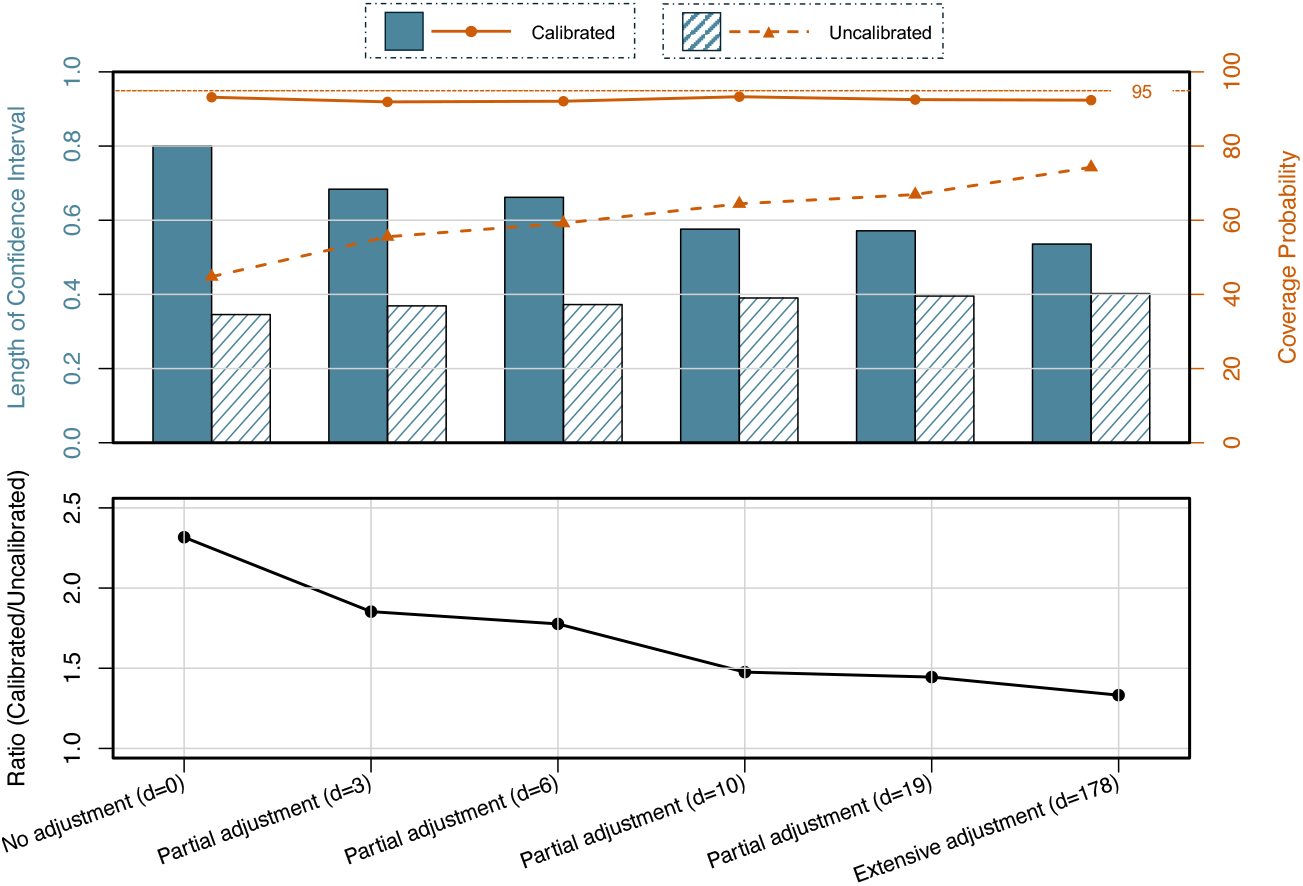
Precision and coverage under DC calibration across covariate adjustment levels. Across covariate adjustment levels, we summarize (a) the mean length of nominal 95% confidence intervals (CIs) before and after LOO-based DC calibration, (b) the corresponding empirical coverage for negative controls, and (c) the ratio of calibrated to uncalibrated CI lengths. Curves and boxplots summarize results across 200 random subsamples.

These results highlight a complementary relationship between adjustment and calibration. Covariate adjustment reduces residual systematic error and shrinks the dispersion of negative-control estimates, which in turn lowers the uncertainty that DC must propagate during calibration. DC then accounts for the remaining systematic error and yields calibrated inference, with the efficiency of the final estimates improving as pre-calibration adjustment becomes stronger.

### Comparative effectiveness across cardiovascular, mental health, and genitourinary outcomes

GLP-1RAs and SGLT2is are widely used glucose-lowering therapies for individuals with type 2 diabetes, and both have established cardiometabolic benefits in randomized trials. To characterize their comparative outcome profiles across multiple clinically relevant domains, we applied DC calibration to 15 incident outcomes spanning cardiovascular (acute hemorrhagic cerebrovascular disease, acute phlebitis/thrombophlebitis and thromboembolism, acute pulmonary embolism, cerebral infarction, hypotension, nonspecific chest pain), mental health (alcohol-related disorders, anxiety and fear-related disorders, depressive disorders, neurodevelopmental disorders, opioid-related disorders), and genitourinary conditions (acute and unspecified renal failure, chronic kidney disease, erectile dysfunction, menstrual disorders). Each outcome was defined using a 12-month washout period to capture new-onset events.

For effect estimation, we used propensity score stratification with the full set of prespecified covariates (extensive covariate adjustment), followed by DC calibration using the prespecified negative-control panel. Figure 5 summarizes the calibrated log risk ratios (logRRs) and 95% confidence intervals for all 15 outcomes. Across cardiovascular endpoints, calibrated estimates were generally close to the null and confidence intervals included no clear differences between GLP-1RAs and SGLT2is. For example, the calibrated logRR for acute pulmonary embolism was − 0.21 (95% CI, − 0.86 to 0.44), consistent with similar risk between therapies in this setting.

**Figure 5.**
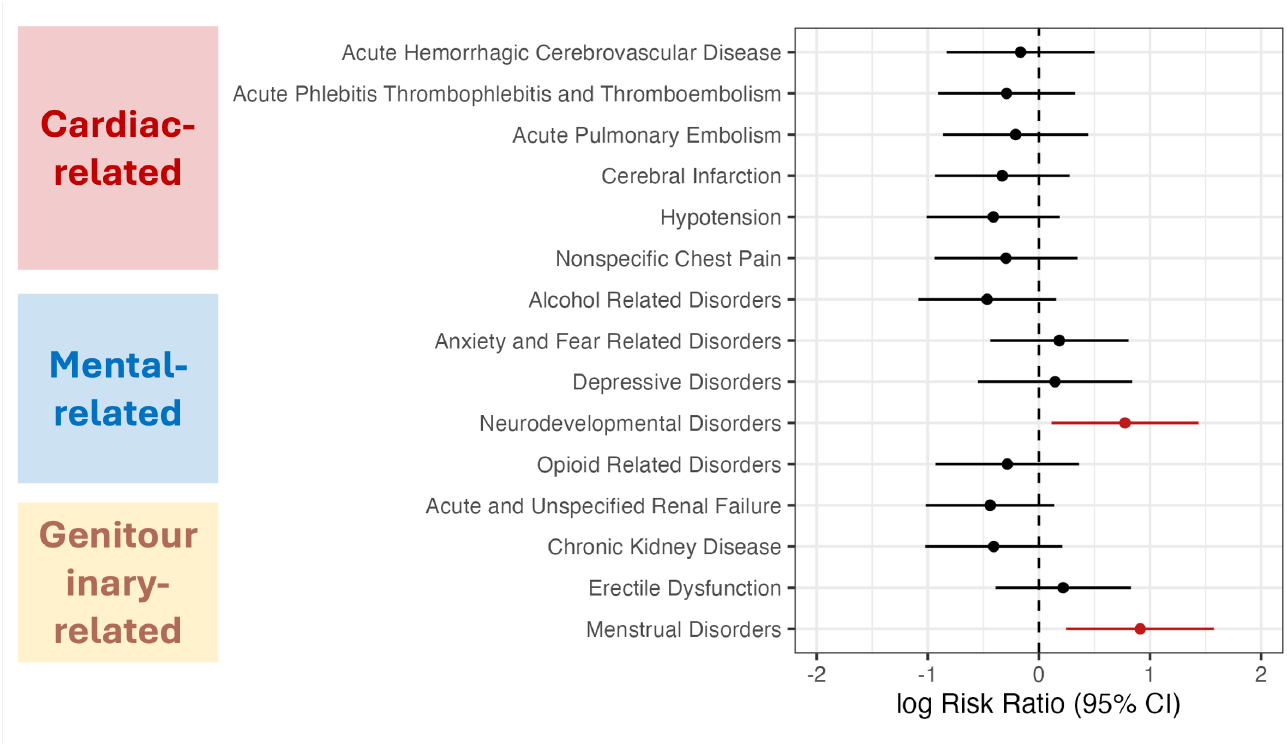
DC-calibrated comparative estimates across 15 prespecified incident outcomes. Shown are calibrated log risk ratios (logRRs) and 95% confidence intervals comparing GLP-1RAs with SGLT2is across cardiovascular, mental health, and genitourinary domains. Red points indicate outcomes for which the calibrated 95% confidence interval excludes the null (logRR = 0).

Mental health outcomes showed greater heterogeneity and wider uncertainty. Most calibrated estimates remained imprecise and crossed the null, indicating limited comparative evidence across these domains in the available data. Neurodevelopmental disorders showed an elevated calibrated estimate for GLP-1RA initiation (logRR 0.77; 95% CI, 0.11 to 1.44); this finding should be interpreted cautiously given the low event rate and the possibility of residual systematic error that may differ across outcome definitions and subpopulations.

For genitourinary outcomes, menstrual disorders showed a positive calibrated association (logRR 0.91; 95% CI, 0.25 to 1.58), whereas erectile dysfunction had a positive point estimate with substantial uncertainty (logRR 0.22; 95% CI, − 0.39 to 0.83).

Overall, these DC-calibrated results provide a bias-aware summary of comparative risks across the prespecified outcome set by propagating empirically observed systematic error from negative controls into uncertainty for the clinical outcomes. This calibration supports more transparent interpretation of outcome-wide real-world comparative effectiveness analyses.

## Discussion

We developed distributional diagnosis and calibration, a framework that uses panels of negative control outcomes to diagnose and calibrate residual systematic error in real-world comparative effectiveness analyses. In the empirical evaluation on a large-scale EHR dataset, DC diagnostics indicated that commonly used adjustment strategies can leave substantial residual systematic error, as reflected by departures from expected null behavior and under-coverage of nominal confidence intervals, particularly under limited covariate adjustment. Applying DC calibration improved alignment with null behavior (e.g., *p*-value uniformity and empirical coverage) across estimation methods. We also found that richer covariate adjustment improved the efficiency of calibrated inference by shrinking the dispersion of NCO effects, thereby reducing the amount of uncertainty that DC must propagate. Together, these results suggest that covariate adjustment and calibration play complementary roles: adjustment reduces systematic error at its source, and DC provides an internal, data-driven safeguard when residual error persists.

A key feature of DC is its *flexibility and modularity*. Because it operates on point estimates and standard errors, DC can be wrapped around a wide range of causal inference pipelines without altering the underlying estimation procedure. This design supports interpretability and reproducibility and makes DC broadly applicable to non-interventional study designs, including observational cohort and case-control studies, self-controlled case series (SCCS), self-controlled risk interval (SCRI) designs, and case-crossover analyses.

DC is particularly relevant for high-stakes settings in regulatory science, pharmacoepidemiology, and public health surveillance, where decisions rely on credible uncertainty quantification and where spurious signals can have substantial downstream consequences [13, 14]. By calibrating inference using an internal empirical null constructed from negative controls, DC emphasizes reliable uncertainty assessment. In practice, associations that remain stable after calibration may be viewed as more robust, whereas estimates that shift toward the null or acquire substantially wider uncertainty warrant more cautious interpretation. More broadly, DC provides a transparent and scalable approach to strengthen real-world evidence pipelines when residual systematic error is difficult to rule out.

Several limitations warrant further study. First, our current calibration models the distribution of NCO-based bias estimates using a parametric form. While this approximation is supported empirically in our analyses and can be justified under mild conditions (Supplementary Information), more flexible or assumption-lean calibration strategies may further improve robustness. Second, DC depends on the validity and representativeness of the NCO panel: violations such as latent causal pathways or outcome misclassification could degrade performance, motivating robust procedures that detect and downweight potentially invalid controls. Third, constructing high-quality NCO panels currently relies on domain expertise; future work could integrate semi-automated candidate generation using biomedical knowledge resources (e.g., SemMedDB [15].) with clinical review to improve scalability in high-throughput applications.

Overall, DC operationalizes a general principle: leverage prior causal knowledge encoded by negative controls to learn about systematic error affecting outcomes of interest. This complements existing causal pipelines by enabling data-driven diagnosis and calibrated uncertainty without requiring explicit modeling of the full data-generating process. We view DC as a practical step toward improving the reliability and interpretability of outcome-wide real-world evidence, particularly in settings where rigorous error control is essential.

## Methods

### Data source and cohort construction

We used data from the TriNetX Research Collaborative Network. This network provides access to de-identified, longitudinal electronic medical records from approximately 152.7 million patients across 106 healthcare organizations, including detailed information on diagnoses, procedures, medications, laboratory values, and genomic data. The data are accessible via the TriNetX Analytics Network (https://trinetx.com). The use of TriNetX for this study was reviewed and approved by the *Institutional Review Board* of the University of Pennsylvania #858496.

#### Target clinical question and outcome

Our clinical objective was to evaluate the comparative effectiveness of GLP-1RAs versus SGLT2is of cardiovascular, mental health, and genitourinary outcomes. Detailed eligibility criteria are provided in *Methods*, and the specific medications included in each treatment group are listed in the *Supplementary Materials*.

#### Eligibility criteria

The study period spanned from January 1, 2019 to September 30, 2024. We included individuals aged 18 years or older who initiated a GLP-1RA or SGLT2i during this period, with no prescriptions for either medication in the previous 12 months. The index date was defined as the date of first prescription for either drug. Patients were required to have at least one healthcare encounter (outpatient, inpatient, or emergency department) in the 12 months prior to the index date. Those who initiated both treatments on the same day were excluded. Participants with specific outcomes during the baseline period (12 month prior to index date) were excluded.

#### NCO selection

A pre-specified list of 95 NCOs, adapted from existing literature [16], was used to evaluate and correct for residual bias and other sources of systematic error. Details can be found in *Supplementary Materials*.

#### Covariate selection

Patient covariates were selected to account for potential confounding which included demographic variables (age, sex, race/ethnicity), clinical characteristics (e.g., baseline comorbidities such as hypertension, obesity, and type 2 diabetes mellitus (T2DM)), and year of cohort entry (ranging from January 2019 to September 2024). These variables were treated as confounders and selected based on clinical relevance and prior literature. However, it should be noted that, as a key challenge in EHR-based studies, not all relevant confounders are routinely or reliably captured, making the presence of unmeasured confounding a persistent concern.

### Causal estimation methods deployment

#### Estimation methods

To adjust for measured confounding, we employed four commonly used estimation methods: propensity score (PS) matching, PS stratification, inverse probability of treatment weighting (IPTW), and multivariable regression with direct covariate adjustment. These approaches enable a comparison of calibration performance across different adjustment paradigms.

#### Adjustment levels

To assess how covariate inclusion affects bias calibration, we defined three levels of adjustment, where *d* is the number of covariates included: (1) no adjustment, with no covariates included (*d* = 0); (2) partial adjustment, using nested subsets of clinically relevant covariates (*d* = 3, 6, 10, 19); and (3) extensive adjustment, incorporating all available demographic and clinical covariates (*d* = 178). Further details are provided in *Methods*.

#### Evaluation dimensions

We evaluated the impact of different covariate adjustment strategies and the proposed distributional calibration procedure across three dimensions: diagnostic performance before and after calibration, and estimation efficiency.

#### Causal estimation methods

To mitigate the effects of observed confounders, we applied four commonly used methods: propensity score (PS) matching, PS stratification, PS inverse probability of treatment weighting (IPTW), and direct covariate adjustment via regression.

The first three methods rely on a two-step procedure. In the first step, we estimated the propensity score using logistic linear regression, modeling the probability of receiving treatment *A* as a function of the observed covariates *X*. In the second step, we estimated treatment effects using outcome regression (*Y* ~ *A*) after adjusting the observed covariates by each method using the estimated propensity scores. A modified Poisson linear regression model [17] was used due to its favorable properties for estimating risk ratios [18].

For PS matching, individuals in the treatment and control groups were matched in a 1:1 ratio, and the outcome model was applied to the matched cohort. For PS stratification, individuals were grouped into quintiles of the estimated PS, and a conditional Poisson regression adjusting for the PS strata was used. For PS weighting, we used stabilized weights based on the inverse of the estimated PS (for treated) or one minus the PS (for controls), and we estimated treatment effects via a weighted Poisson regression. Lastly, for outcome regression adjustment, we directly fit a multivariate modified Poisson regression model including all observed covariates.

#### Selection of partially adjusted covariates

To evaluate the robustness of distributional calibration under varying levels of confounding adjustment, we predefined four nested tiers of partial covariate adjustment. Each tier incrementally incorporated clinically meaningful variables selected based on domain expertise and prior literature, reflecting potential confounders of the treatment-outcome relationship.

- Demographics only (3 covariates): Basic demographic covariates, including age, sex, and race/ethnicity.
- Demographics and Core Metabolic Comorbidities (6 covariates): Demographics plus key metabolic comorbidities, including age, sex, and race/ethnicity, type 2 diabetes mellitus, obesity, and hypertension.
- Core and Cardiovascular Conditions (10 covariates): All covariates above, plus additional cardiometabolic comorbidities, including disorders of lipid metabolism, heart failure, cardiac arrhythmias, cardiorespiratory signs and symptoms
- More Clinical Profile (19 covariates): The most comprehensive partial set, further including sleep-wake disorders, anxiety disorder, general signs and symptoms, chest pain, chronic kidney disease, abdominal pain, low back pain, headache, and asthma.

The extensive adjustment list included 178 covariates, encompassing demographics (age, sex, race/ethnicity), detailed clinical characteristics, and the calendar year of cohort entry (ranging from January 2019 to September 2024). The codeset for clinical conditions is available at https://github.com/Penncil/Distributional-diagnosis-and-calibration/blob/master/cluster_master.csv, and conditions with a prevalence below 0.5% were excluded to enhance computational efficiency.

### Statistical notation

We first introduce necessary notation. We adopt the potential outcomes framework [19], where *A* ∈ {0, 1} denotes a binary treatment, *X* the observed covariates, and *U* unmeasured confounders. For each outcome *Y*_*j*_, let *Y*_*j*_(1) and *Y*_*j*_(0) denote the potential outcomes under treatment and control, respectively, with *Y*_*j*_ = *AY*_*j*_(1) + (1 −*A*)*Y*_*j*_(0). We index the primary outcome as *Y*_0_, and define *Y*_*j*_ with *j* ≥ 1 as an NCO if E {*Y*_*j*_(1)} = E {*Y*_*j*_(0)}. We observe *n* independently and identically distributed (i.i.d.) samples of (*Y*_i,0_, *A*_i_, *X*_i_, *Y*_i,1_, …, *Y*_i,J_), *i* = 1, …, *n*. See Figure 1 for illustration.

Let *β*_*A*_ denote a generic notation of the target causal estimand for the primary outcome *Y*. The causal parameter *β*_*A*_ may vary depending on context and application. Common choices include:

- **The average treatment effect**: *β*_ATE_ := E{*Y* (1) − *Y* (0)};
- **The log risk ratio**: *β*_logRR_ := log {E{*Y* (1)}*/*E{*Y* (0)}}.

For a given estimation method of *β*_*A*_, let 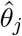 denote the estimated treatment effect for the *j*th outcome/NCO, where *j* = 0 corresponds to the outcome of interest and *j* = 1, …, *J* index a set of NCOs. Let 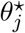 denote the asymptotic limit of 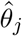, which represents the value of the estimate by the estimation procedure as if we had infinitely many samples. Note that 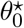 may not equal *β*_*A*_ and 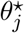 may not equal 0 for NCOs due to systematic error. In the main text, we focus on the causal parameter ATE, namely *β*_*A*_ = *β*_ATE_, though the methodology generalizes to other estimands.

### Framework overview: distributional diagnosis and calibration with NCOs

Importantly, the framework does not require specification of a data-generating model or causal directed acyclic graphs, making it broadly applicable across settings. See *Supplementary Materials* (Examples S.1–S.3) for detailed discussions of its use under different structural assumptions.

### DC framework layer (a) — setup stage: integrating NCOs into the causal estimation pipeline

In comparative effectiveness research, it is a standard practice to follow a structured protocol: define the scientific question, select the eligible cohort, identify relevant covariates, and estimate treatment effects using adjustment strategies such as propensity score matching. Our framework extends this protocol by incorporating NCOs for bias diagnosis and correction. The setup proceeds as follows:

1. *Define the causal question and eligible population*. Specify the treatment variable *A* and primary outcome *Y*_0_ based on the causal estimand of interest. Define the eligible population from the target real-world database using prespecified inclusion and exclusion criteria.
2. *Select key covariates and identify NCOs*. Use prior scientific and clinical knowledge to identify potential confounders and NCOs. Specifically, NCOs are outcomes *Y*_1_, …, *Y*_*j*_ that are known to be causally unaffected by *A* but share a similar confounding structure with the primary outcome.
3. *Estimate treatment effects*. Apply the chosen estimation method to each outcome to obtain a collection of point estimates and associated variance estimates: 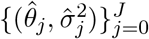, where *j* = 0 corresponds to the primary outcome and *j* = 1, …, *J* index the NCOs.

This setup stage establishes the foundation for the subsequent diagnostic and calibration procedures. A schematic illustration of this setup is provided in panel (a) of Figure 1.

### DC framework layer (b) — diagnostic layer: detecting residual bias

We illustrate the proposed diagnosis method in panel (b) of Figure 1. Given an estimation procedure, we assess whether it sufficiently removes the residual confounding bias using the estimates 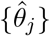 and standard errors 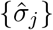 from the NCOs. We propose two complementary diagnostic strategies: (i) the uniformity deviation area test, and (ii) empirical negative control coverage evaluation.

#### Quantile-quantile plot and uniformity deviation area

Under the null hypothesis that the estimator is unbiased, treatment effects on NCOs should be centered at zero, and the corresponding *p*-values should follow a Uniform(0, 1) distribution. We visualize this behavior using Quantile-Quantile (QQ) plots and summarize deviations from the uniformity line (the 45-degree diagonal) by the total area between the QQ curve and the diagonal. We term this quantity the *Uniformity Deviation Area* (UDA), which serves as a scalar diagnostic for systematic bias. Formally, UDA can be used to test the hypothesis:

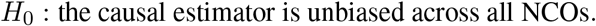

Using the parametric bootstrap, we derive the empirical distribution of UDA under *H*_0_ in *Methods*, enabling principled statistical inference. Since the *p*-values for NCOs are computed on the same dataset, they are generally correlated, violating the independence assumptions required by classical asymptotic theory. This dependence structure motivates the use of the parametric bootstrap, which accounts for such correlations and yields more accurate empirical quantiles. In practice, we estimate the 95th and 99th percentiles of this null distribution using *B* = 1,000 bootstrap replicates to ensure stable quantile estimation. If the observed UDA falls below these critical values, we retain *H*_0_ and interpret the result as consistent with unbiased estimation. Conversely, if the observed UDA exceeds these thresholds, we have statistical evidence to reject *H*_0_, suggesting the presence of systematic bias in the estimation procedure.

#### Empirical coverage probability

As a complementary diagnostic, we evaluate the empirical coverage of the confidence intervals constructed for NCOs. Since the true treatment effect on each NCO is known to be zero based on prior knowledge, correct coverage requires that zero falls within the nominal (1 − *α*) confidence interval at the expected frequency. For each NCO, we record whether its confidence interval covers zero, and average across *J* outcomes to estimate empirical coverage. We refer to this procedure as the *Empirical negative control coverage* (ENCC).

Since the empirical coverage is itself a random quantity based on a finite number of NCOs, it naturally fluctuates around the nominal level. For instance, with *J* NCOs and a nominal level of 1 − *α* = 0.95, the empirical coverage under the null follows a Binomial distribution Bin(*J*, 0.95). Under the central limit theorem, an approximate 95% confidence interval for the empirical coverage can be computed as [0.95 ± 1.96 {0. 95 × 0.05*J* ^−1^}^1/2^], which evaluates to approximately [0.91, 0.99] when *J* = 95. A significant drop below this range suggests residual bias or misestimated variance.

### DC framework layer (c)—calibration layer: correcting for residual bias

Panel (c) of Figure 1 is a schematic illustration of the distributional calibration. The diagnostic and calibration layers are implemented as a unified two-stage procedure. In particular, when the diagnostics suggest substantial bias, the calibration has a greater effect, whereas when residual bias is negligible, the calibration leaves the primary estimate largely unchanged. This is done by modeling the distribution of biases implied by the NCO estimates. Specifically, we compute 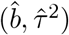, where 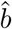 denotes the average estimate of NCO causal effects, and 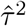 denotes the variance of estimated NCO causal effects. These quantities are then used to construct a bias-corrected estimator and adjusted variance for the primary outcome:

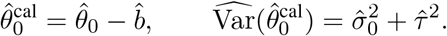

The resulting calibrated confidence interval is given by:

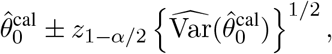

which is provably valid under mild regularity conditions. Here, *z*_1−α/2_ denotes the (1 − *α/*2) standard normal quantile. Further technical details are provided in *Methods*.

#### LOO-based DC: evaluating DC through the leave-one-out strategy

In the real-world example, the same set of NCOs is used for both diagnosis and calibration. We implement a leave-one-out (LOO) strategy to prevent circular inference. Specifically, in each iteration, one NCO is held out, and the remaining *J* − 1 NCOs are used to estimate the residual bias distribution. Let 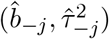 denote the estimated mean and variance of residual bias computed from the *J* − 1 NCOs excluding the *j*th one. Here, 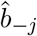 captures the average residual bias and 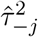 reflects its variability. These quantities are then used to calibrate the treatment effect estimate and variance for the held-out NCO:

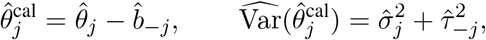

where 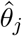 denotes the point estimate for the *j*th NCO produced by the chosen estimation method, and 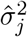 is its corresponding model-based variance estimate. We refer to this procedure as the LOO-powered distributional calibration.

We then apply the UDA and ENCC diagnostics to the cross-validated collection 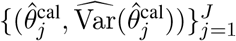. Specifically, these calibrated estimates are used to compute *p*-values for uniformity assessment and to evaluate empirical coverage after calibration. This design ensures that diagnostic evaluations reflect the performance of the bias-corrected procedure itself, free from circularity or self-referential inflation. We refer to this diagnostic procedure as the LOO-based DC.

### Framework for DC

Due to unmeasured confounding, the asymptotic estimand 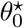 by the chosen estimation method for the outcome of interest may differ from *β*_*A*_, and we write

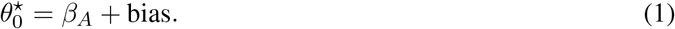

Since *β*_*A*_ is not directly identifiable from observed data, we leverage NCOs to infer the residual bias. The key idea is that the true causal effect for each NCO is zero; hence, their asymptotic estimates 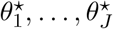 reflect residual biases due to unmeasured confounders rather than causal relationships. Formally, we make the following assumption for identification of *β*_*A*_.

#### Assumption 1

*The asymptotic biases for NCOs follow a common distribution independently*,

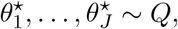

*and that the residual bias in the primary outcome is exchangeable with those of the NCOs:* 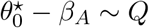. *Here, Q represents a latent distribution that governs the bias structure across outcomes*.

Assumption 1 formalizes the idea of distributional exchangeability, stating that the residual bias in the target outcome is drawn from the same distribution as that of the NCOs. While this assumption is not empirically verifiable, they can be justified by design: for instance, when NCOs are selected to mirror the confounding structure of the outcome of interest, their bias distributions are expected to follow a common generative process. In *Supplementary Materials*, we give interpretations for *Q* under linear models. Moreover, the assumption of distributional exchangeability is notably flexible. It does not require the underlying confounding mechanisms to be identical across outcomes, only that the induced biases are governed by a shared probabilistic model. This mild assumption enables the method to accommodate diverse forms of causal directed acyclic graphs (DAG) commonly in real-world settings. See Examples S.1–S.3 for more details.

### Diagnosis for residual bias: uniformity area deviation test

To formally assess whether a candidate estimation method sufficiently removes residual confounding, we introduce a nonparametric goodness-of-fit test based on the empirical distribution of *p*-values from NCOs. The diagnostic statistic, termed UDA, quantifies the *L*_1_ distance between the empirical quantiles of observed *p*-values and the theoretical quantiles of the Uniform(0, 1) distribution.

For a given estimation method, if it were unbiased, regular, and asymptotic linear, the standardized treatment effect estimate for each NCO should satisfy 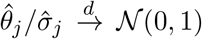. Accordingly, we compute a two-sided *p*-value for each NCO as

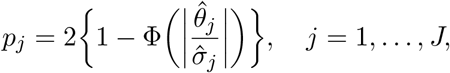

where Φ(·) denotes the cumulative distribution function of the standard normal distribution. These *p*-values are then used to assess the unbiasedness of the estimation method via uniformity-based diagnostics. Here, 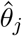 is the point estimate produced by a given estimation procedure, and 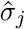 is the associated standard deviation. This framework applies not only to conventional estimators but also to the LOO-powered calibrated estimator introduced in our method.

Let *C*(*t*) denote the empirical quantile function of the *p*-values, as plotted in the QQ plot. We define the uniformity deviation area as the area between the empirical QQ curve and the identity line *y* = *t*, given by the integral

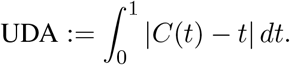

In practice, we approximate this area via a simple discretization. Let *p*_(1)_ ≤ *p*_(2)_ ≤…≤ *p*_(J)_ denote the order statistics of the *p*-values. The UDA test statistic is computed as

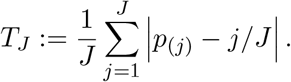

A large value of *T*_*j*_ suggests systematic deviation from uniformity and thus the estimation method is biased. In the *Supplementary Materials*, we compute the quantiles of the UDA statistic *T*_*j*_ under the null using a parametric bootstrap procedure. Here, we illustrate the procedure in the context of propensity score matching.

First, we follow the standard workflow to estimate the propensity score, perform matching, and compute the treatment effect estimates along with the resulting UDA statistic *T*_*j*_. Second, for each bootstrap iteration indexed by *b, b* = 1, …, *B*, we generate a new treatment vector 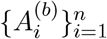 by sampling from Bernoulli 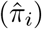, where 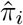 denotes the estimated propensity score for unit *i*. Finally, using the resampled treatment assignments 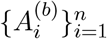, we re-estimate the propensity score, apply the matching procedure, and recompute 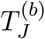. Because the resampled treatment assignments depend only on the observed covariates, the resulting estimates are not subject to residual confounding bias. Repeating this procedure for *B* iterations yields an empirical distribution of 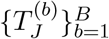 that approximates the distribution of *T*_*j*_ under the null. This process enables us to compute critical values for *T*_*j*_ without relying on large-sample approximations.

### Distributional set identification result

Under Assumption 1, the central (1 − *α*) quantile interval of *Q* directly yields a confidence region for the residual bias in the target estimand. That is, denoting by *q*_α/2_ and *q*_1−α/2_ the *α/*2 and 1− *α/*2 quantiles of *Q*, then

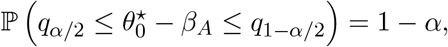

implying that *β*_*A*_ is contained within 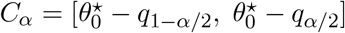 with confidence level 1 − *α*. We now state a formal condition under which confidence intervals for the target causal effect *β*_*A*_ are identified via the distributional calibration framework.

#### Proposition 1

*Suppose that Assumption 1 holds. Then a* (1−*α*) *confidence interval* 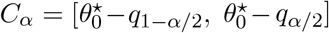 *for β*_*A*_ *can be identified such that* P(*β*_*A*_ ∈ *C*_*α*_) = 1 − *α*.

It is noteworthy that this form of identification departs from classical frameworks in causal inference, for example, point identification, where the target estimand *β*_*A*_ is determined exactly under strong assumptions, including a correctly specified DAG for the causal relationships among variables. Our approach also differs from set identification strategies [20], where a deterministic range of the estimand is obtained; for example, those used in sensitivity analysis and instrumental variable settings [21, 22].

By contrast, our framework yields a distributionally calibrated confidence region that reflects uncertainty in residual bias across multiple NCOs. We refer to this as *distributional set identification*, as it constructs a confidence region for the target estimand based on an estimated bias distribution. Unlike point identification or sensitivity analysis, this approach provides an interval estimate with a coverage guarantee on the true causal effect based on the empirical data. Notably, when the residual bias distribution contracts to a point mass, the calibrated estimator becomes point-identified in the asymptotic limit; see the discussion following Theorem 1 for details.

In practice, we estimate 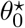 by 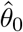, and infer quantiles of the distribution *Q* from the NCO estimates 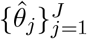. In our implementation, we focus on the normal case *Q* = *N*(*b, τ* ^2^), for which the parameters (*b, τ* ^2^) can be consistently estimated using method-of-moments estimators (see (2)). Nevertheless, the framework is not limited to the Gaussian case. More generally, the distribution *Q* can be estimated nonparametrically using techniques from empirical Bayes, such as the nonparametric maximum likelihood estimator (NPMLE) for a mixing distribution [23, 24]. These approaches allow inference on the quantiles of *Q* without imposing strong parametric assumptions, and can be readily incorporated into our calibration procedure.

### Distributional calibration algorithm

For clarity, we separate the workflow into three transparent steps, which can be implemented with a few lines of code.

#### Step 1: Adjustment for observed confounders

Given an estimation method of *β*_*A*_ to adjust for observed confounders, such as outcome regression, inverse probability weighting/matching, or augmented inverse probability weighting, apply it to the outcome of primary interest and NCOs in the study cohort. Obtain the estimate 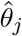 and its standard deviation 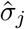 for the outcome (*j* = 0) and each NCO (*j* = 1, …, *J*).

#### Step 2: Estimation for unobserved confounder biases

Assume normal distribution *Q* = *N* (*b, τ* ^2^) for the estimation biases, the method-of-moments estimators of (*b, τ* ^2^) are

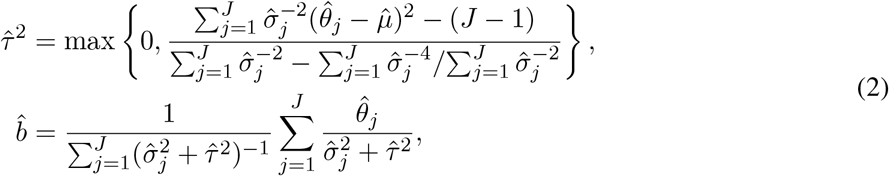

where 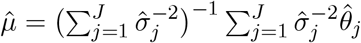 denotes the fixed-effect meta estimator for *b*. Here, 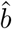 can be viewed as the marginal mean of the bias effect.

#### Step 3: Distributional calibration

The calibrated estimate of *β*_*A*_ and its variance for the outcome of interest are

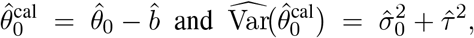

yielding a 100(1 − *α*)% confidence interval 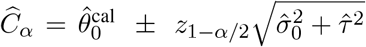 where *z*_1−*α*/2_ denotes the (1 − *α*/2) quantile of the standard normal distribution.

### Theoretical guarantees of DC

The following results show that the estimate of the bias distribution *Q* is consistent, and the confidence interval Ĉ_α_ achieves asymptotic coverage. Those results imply a valid inference of the target causal effect *β*_*A*_ by the proposed DC approach in the presence of unmeasured confounders.

#### **Theorem 1** (Asymptotic coverage)

*Suppose that Assumption 1 holds with Q* = *N* (*b, τ* ^2^). *If the regularity condition S*.*1 in the* Supplementary Materials *holds, then*

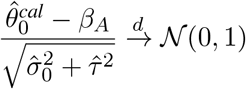

*as n, J* → ∞. *Consequently, the Wald-type confidence interval Ĉα achieves asymptotic* (1 − *α*) *coverage for β*_*A*._

As discussed in Proposition 1, one can only identify a valid (1 −*α*) confidence interval for *β*_*A*_; in this case, the calibrated point estimate 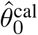 is not meaningful as a consistent estimator. However, when the residual bias variance diminishes asymptotically, e.g., *τ* → 0, the proof of Proposition S.2 in the *Supplementary Materials* also implies that 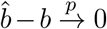, 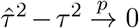. In this regime, the identified confidence interval contracts to a point as the sample size grows, and the calibrated estimate 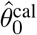 becomes a consistent estimator for *β*_*A*_.

These results provide theoretical justification for the DC procedure as a valid bias correction layer. Notably, they demonstrate that the utility of DC does not hinge on full identifiability of the causal effect or the complete removal of confounding. Rather, it is sufficient to consistently estimate the distribution of residual bias across a collection of NCOs, enabling valid inference under partial confounding adjustment.

### DC is agnostic to estimation methods and benefits from stronger covariate adjustment

In this section, we investigate two key problem of DC using linear models:

i. Under what structural assumptions on the data generating mechanism do the biases of the outcome of interest and NCOs follow a common distribution, given a set of covariates and a pre-specified estimation method?
ii. How does the level of covariate adjustment influence the efficiency and statistical power of the calibrated estimator?

To answer the two questions, we consider the following linear data-generating model:

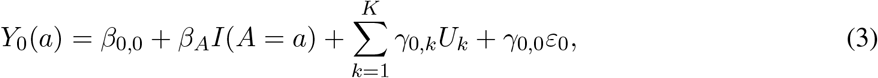

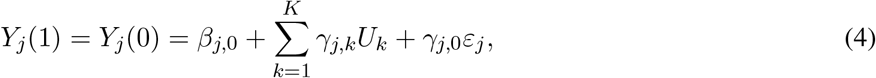

where *U*_1:*K*_ := (*U*_1_, …, *U*_*K*_) denote *K* unmeasured confounders, and *ε*_*j*_ are independent additive noise terms for each outcome. The binary treatment *A* ∈ {0, 1} depends on *U*_1_, …, *U*_*K*_, and the noise terms *ε*_*j*_ are independent of (*A, U*_1:*K*_), with *ε*_*j*_ ~ *N*(0, 1). The observed outcome is defined as *Y*_0_ = *AY*_0_(1) + (1 − *A*) *Y*_0_(0) and *Y*_*j*_ = *AY*_*j*_(1) + (1 − *A*)*Y*_*j*_(0) = *Y*_*j*_(1) = *Y*_*j*_(0). Let *P*_*j*_ be the joint distribution of (*A, Y*_*j*_, *U*_1:*K*_) and *P*_*j,t*_ the joint distribution of (*A, Y*_*j*_, *U*_1:*t*_).

To better understand the role of covariate adjustment in distributional calibration, we consider a setting in which the first *t* confounders, denoted by *U*_1:*t*_, are assumed to be observed. That is, we observe *n* independent observations (*Y*_*i*,0_, *Y*_*i*,1_, …, *Y*_*i,J*_, *A*_*i*_, *U*_i,1:*t*_) from the model in (3) and (4) for *i* = 1, …, *n*. This setup allows us to explore how adjusting varying levels of observed confounders influences both the distribution of estimation bias and the efficiency of the calibrated estimator.

#### Estimation agnosticism of DC

We argue that the validity of the proposed DC procedure does not depend on the specific choice of the estimation method. To elucidate the estimation-agnostic nature of DC, we consider a general class of estimation procedures. Let 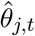 denote the point estimate of the treatment effect for outcome *j*, obtained by applying a chosen estimation procedure to the observed data 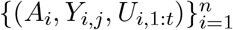. Define 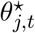 as the probability limit of 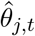 as *n* → ∞. It should be noted that some restrictions of estimation procedures should be made: procedures that, for example, arbitrarily assigning values for the estimates of NCOs would violate Assumption 1 and are excluded from consideration.

In this paper, we assume that the asymptotic limit of the treatment effect estimator for the outcome of interest takes the form

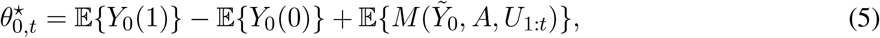

where 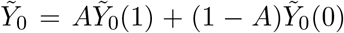 with 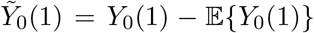 and 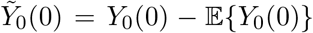, and *M*(·) represents some function uniquely determined by the estimation procedure. (5) holds if the estimating procedure can produce asymptotically unbiased estimates for the causal estimand when there are no unmeasured confounders.

For the observed NCOs, the causal effect of treatment is zero by design. Thus, the corresponding asymptotic limit takes the form

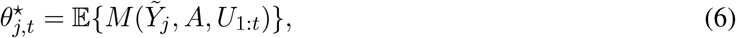

where 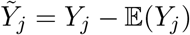 denotes the centered version of the *j*th NCO. This formulation in (5) enables direct comparison of residual biases across outcomes on a common, mean-zero scale. Moreover, a variety of estimation procedures satisfy satisfy (5) and (6); for example, we show in the *Supplementary Materials* that the inverse propensity score weighting estimation and the outcome regression estimation meet this assumption.

##### Theorem 2.

*Suppose the data are generated from the linear model specified in equations* (3)*–*(4), *and that the first t confounders U*_1:*t*_ *are observed. Assume the following conditions hold:*

- *Confounders are mean zero, i*.*e*., E(*U*_*K*_) = 0 *for k* = 1, …, *K;*
- (5) *and* (6) *hold for some unknown function M* (·) *uniquely determined by the estimation procedure;*
- *The coefficients* (*γ*_*j*,0_, …, *γ*_*j,K*_)^⊤^ *are independently drawn across outcomes j from a shared distribution*.

*Then, the limiting quantities* 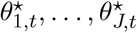 *and* 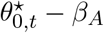 *are exchangeable and follow a common distribution determined by the law of* (*γ*_j,0_, …, *γ*_j,*K*_)^⊤^.

This result formalizes the key assumption underlying the proposed DC framework: namely, after adjusting for the observed confounders *U*_1:*t*_, the residual biases across outcomes, quantified by the limiting estimands 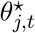, are exchangeable and follow a shared distribution. Importantly, this property arises directly from the structure of the data-generating model instead of the specific estimation method.

#### Efficiency gain from stronger covariate adjustment

In what follows, we provide two examples, the propensity score weighting estimator and the outcome regression estimator, to illustrate that stronger covariate adjustment before DC can improve efficiency. Specifically, under inverse probability weighting (IPW), the estimated treatment effect for the *j*th outcome is

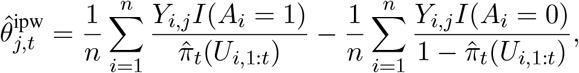

where 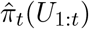 is an estimator for the propensity score *π*_*t*_(*U*_1:*t*_) = P(*A* = 1 | *U*_1:*t*_), and denote by 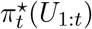 the probabilistic limit of 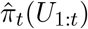. Then,

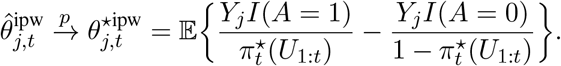

Alternatively, using outcome regression, the estimated treatment effect is

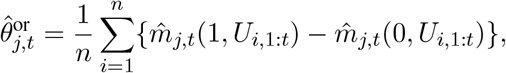

where 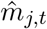 is an estimator for *m*_j,*t*_(*a, U*_1:*t*_) = E(*Y*_*j*_ | *A* = *a, U*_1:*t*_), and denote by 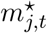 the probabilistic limit of 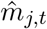 Then,

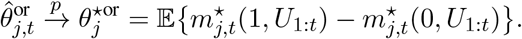

Next, we will show that when working models are correctly specified, the variability of the resultant asymptotic limit decreases as the adjustment level become stronger. Before stating the result, we define *δ*_*K*_(*u*_1:*t*_) = E(*U*_*K*_ | *A* = 1, *U*_1:*t*_ = *u*_1:*t*_) − E(*U*_*K*_ | *A* = 0, *U*_1:*t*_ = *u*_1:*t*_).

##### Proposition 2.

*Suppose that the data generating model* (3) *and* (4) *holds. If* 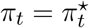 *and* 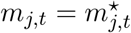, *then* (5) *and* (6) *hold with*

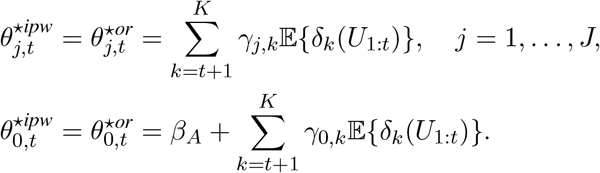

These structured forms in Proposition 2 provide two insights. First, as shown in Proposition S.2 of the *Supplementary Material*, when the number of unmeasured confounders *K* is large, the assumption that each *γ*_*j,k*_ follows the same distribution across outcomes *j* can be relaxed. In this high-dimensional regime, it suffices that the *weighted sums* of the means and variances of *γ*_*j,k*_ converge to common constants across *j*. Under this condition, the residual biases remain approximately exchangeable, thereby justifying the use of distributional calibration. Second, as the number of observed confounders *t* increases, the number of unadjusted terms contributing to the residual bias decreases. This implies a reduction in the variability of the bias across outcomes, meaning that the uncertainty introduced by calibration diminishes. This property is naturally captured by the NCO calibration framework, which reflects the efficiency gain from stronger covariate adjustment.

## Data availability

This study used population-level aggregate and de-identified data generated by the TriNetX platform. Due to data privacy, patient-level data cannot be shared.

## Code availability

To facilitate adoption, we provide an open-source R package, *debiasedTrialEmulation*, available at https://cran.r-project.org/web/packages/debiasedTrialEmulation/index.html. While originally developed to support target trial emulation workflows, the package is fully modular: the distributional calibration module can be used independently of the trial emulation components. This allows users to incorporate our bias correction method into a wide range of causal inference pipelines. The package includes functions for fitting standard adjustment methods, estimating bias distributions from NCOs, and generating calibrated confidence intervals. Comprehensive documentation and example workflows are provided to support integration into large-scale real-world evidence studies.

